# Mass Spectrometry-Based Proteomic Profiling of Sonicate Fluid Differentiates *Staphylococcus aureus* Periprosthetic Joint Infection from Non-Infectious Failure: A pilot study

**DOI:** 10.1101/2022.12.28.22284010

**Authors:** Cody R. Fisher, Kiran K. Mangalaparthi, Kerryl E. Greenwood-Quaintance, Matthew P. Abdel, Akhilesh Pandey, Robin Patel

## Abstract

**Purpose:** This study aims to use proteomic profiling of sonicate fluid samples to compare host response during *Staphylococcus aureus*-associated periprosthetic joint infection (PJI) and non-infected arthroplasty failure (NIAF) and investigate novel biomarkers to increase diagnostic accuracy.

**Experimental Design:** In this pilot study, eight sonicate fluid samples (four from NIAF and four from *Staphylococcus aureus* PJI) were studied. Samples were reduced, alkylated and trypsinized overnight, followed by analysis using liquid chromatography-tandem mass spectrometry (LC-MS/MS) on a high-resolution Orbitrap Eclipse mass spectrometer. MaxQuant software suite was used for protein identification, filtering, and label-free quantitation.

**Results:** Principal component analysis of the identified proteins clearly separated *S. aureus* PJI and NIAF samples. Overall, 810 proteins were quantified in any three samples from each group and 35 statistically significant differentially abundant proteins (DAPs) were found (2-sample t-test p-values ≤0.05 and log_2_fold-change values ≥2 or ≤-2). Gene ontology pathway analysis found that microbial defense responses, specifically those related to neutrophil activation, were increased in *S. aureus* PJI compared to NIAF samples.

**Conclusion and Clinical Relevance:** Proteomic profiling of sonicate fluid using LC-MS/MS, alone or in combination with complementary protein analyses, differentiated *S. aureus* PJI and NIAF in this pilot study.

## 1. INTRODUCTION

Periprosthetic joint infection (PJI) is a major cause of arthroplasty failure after total knee or hip replacement surgery. Of the estimated 7 million Americans living with a total knee or hip arthroplasty as of 2010, about 1-2% will go on to get a PJI ^[1-5]^. As the number of individuals who undergo arthroplasty surgery continues to rise, so will the number of PJIs, resulting in extended hospital stays, increased antimicrobial usage, and an estimated $1.85 billion in annual healthcare costs by 2030 ^[6-9]^. *Staphylococcus aureus* is a common cause of PJI, associated with robust biofilm production, production of numerous virulence factors, and resistance to antimicrobial agents and the host immune response ^[1,2,10-15]^.

Some PJI cases can be difficult to differentiate from non-infectious arthroplasty failure (NIAF) ^[16-21]^. Typically, arthroplasty failure is treated with resection of the failed implant and one- or two-stage revision surgery or debridement and implant retention, alongside an extended course of antimicrobial agents ^[1,22-24]^. Microbial- and host-based assays are used to diagnose PJI. Microbial diagnostics may be limited by negative growth in culture or negative results of molecular diagnostics, or conversely, falsely-positive results as a result of detection of contaminants ^[25]^. Additional host-derived biomarkers for PJI, such as erythrocyte sedimentation rate in blood; nucleated cell count and neutrophil percentage in synovial fluid; and intraoperative tissue histology and purulence, can provide evidence of underlying infection ^[1,22,26]^. Host proteomic profiling has also been shown to have the potential to differentiate PJI from NIAF, with increased expression of c-reactive protein, D-dimer, chemokine ligand 20 (CCL20), calprotectin, lipocalin, lactotransferrin, and many others, in PJI compared to NIAF patient samples ^[26-35]^. Measurement of one protein, alpha-defensin, in synovial fluid has been approved by the United States Food and Drug Administration as an aid for the detection of PJI ^[33,36]^.

Multi-omics approaches have recently been investigated as potential alternatives to overcome limitations of currently used diagnostic tools ^[26,29,37-40]^. Previous studies have shown that liquid chromatography-tandem mass spectrometry (LC-MS/MS)-based profiling may be able to differentiate synovial fluid samples from PJI and NIAF patients ^[31,41]^. Analysis of sonicate fluid, a sample-type that directly interrogates the site of infection - the implant surface -, may hypothetically allow for enhanced differentiation of infected and non-infected individuals and potentially PJI biomarker discovery ^[37,40,42-44]^. Mass spectrometry-based proteomic analysis provides an unbiased and in-depth overview of the protein expression alterations in biological samples. Our group has previously employed mass spectrometry-based proteomics in developing assays of potential diagnostic utility ^[45,46]^ and in studying host response to severe acute respiratory syndrome coronavirus 2 (SARS-CoV-2) viral infection ^[47,48]^. This pilot study was aimed to evaluate the feasibility of proteomic analysis of sonicate fluid to reflect the host response to PJI. The advancement of proteomics-based profiling of patient samples during PJI could yield novel insights into underlying biological interactions.

## 2. MATERIAL AND METHODS

### 2.1 Sonicate fluid harvest and cohort

Sonicate fluid samples were collected between October 2013 and July 2017 from patients undergoing revision total hip or knee arthroplasty; arthroplasty components were removed and subjected to sonication for clinical purposes, as previously described; their use in this study was approved by the Mayo Clinic Institutional Review Board (09-000808) (23). Four NIAF and four *S. aureus* PJI sonicate fluid samples were analyzed. Infection status was determined by the 2018 Musculoskeletal Infection Society and International Consensus Meeting criteria ^[49]^.

### 2.2 Protein extraction and LC-MS/MS analysis

100 μl of sonicate fluid samples were adjusted to 0.2% Rapigest followed by heating at 90 °C for 5 minutes. Samples were then reduced with 5 mM tris(2-carboxyethyl)phosphine (TCEP) for 45 minutes at room temperature and alkylated with 20 mM Iodoacetamide for 25 minutes in the dark. Trypsin enzyme was added in the ratio of 1:20 for overnight digestion at 37 °C. Following C_18_ cleanup, 2 μg of peptides were subjected to quantitative LC-MS/MS analysis on a high-resolution Orbitrap Eclipse Tribrid mass spectrometer connected to UltiMate 3000 RSLC nano system (Thermo Scientific, Waltham, MA). Separation of peptides includes initial trapping on a trap column (PepMap C18, 2 cm × 100 μm, 100 Å, Thermo Scientific, San Jose, CA) using 0.1% formic acid (solvent A) followed by gradient elution (3% to 28% to 40%) using acetonitrile, 0.1% formic acid (solvent B) on an analytical column (EasySpray 50 cm × 75 μm, C18 1.9 μm,100 Å, Thermo Scientific, San Jose, CA). Each run was started with equilibration of both columns for 5 minutes and the overall time for each run was 240 minutes. As the peptides were eluting, the Orbitrap was operated in a data dependent mode with a cycle time of 2 seconds. Initially, the precursor MS scan was recorded for 350-1500 m/z using a normalized AGC target of 100%, injection time of 50 ms and 60,000 resolution. Precursors with a minimum intensity threshold of 20,000 and charge states 2-7 were taken for MS/MS starting from high intensity precursors. Monoisotopic precursor selection was also enabled. Quadrupole was used for precursor isolation with 1.6 m/z isolation width and fragmented with normalized HCD energy of 28% and resulting fragment ions was recorded in Orbitrap analyzer. Fragment ion spectrum was recorded at 15,000 resolution using normalized AGC target of 200% and maximum injection time of 120 ms. Dynamic exclusion of 30 seconds was used to prevent repeated fragmentation of the precursor ions.

### 2.3 LC-MS/MS raw data analysis

MaxQuant software suite (V 2.0.1.0) was used for protein identification and quantitation. Database searching was performed against human UniProt protein database with *in silico* trypsin digestion set to be specific and maximum of 2 missed cleavages allowed. Default mass tolerance settings of 20 pm for first search and 4.5 ppm for main search were used. Oxidation (Methionine) and protein N-terminal acetylation were used as dynamic modifications, whereas carbamidomethylation (cysteine) was used as static modification. Proteins were filtered at 1% protein-level false discovery rate (FDR). LFQ algorithm within MaxQuant was used for label-free quantitation and match between the runs option was enabled with match time window of 1 minute to reduce missing values. Finally, label-free quantified values were logarithmized and proteins not quantified in at least three samples in each group were eliminated.

### 2.4 Data organization and statistical analysis

Quantitative data were organized and graphed in RStudio v1.2.5042 ^[50]^ using the R-packages “ComplexHeatmap” ^[51]^ for heatmap creation and “Factoextra” ^[52]^ for principal component analysis. Protein-specific analyses were conducted, and a volcano plot created in Graphpad Prism 9 v9.2.0 (San Diego, CA). Enrichr was used for gene ontology pathway analysis ^[53-55]^. Statistical significance was determined using 2-sample t-test with multiple hypothesis correction (p-values ≤0.05 and log_2_fold-change values ≥2 or ≤-2).

## 3. RESULTS AND DISCUSSION

### 3.1 Cohort differential protein abundance characterization

In this study, quantitative LC-MS/MS analysis was used to characterize the host proteomic profile of four NIAF and four *S. aureus* PJI sonicate fluid samples and determine whether NIAF and *S. aureus* PJI-associated samples can be differentiated. Eight hundred and ten proteins were quantified in at least three of four samples each of *S. aureus* PJI and NIAF cases, of which 35 differentially abundant proteins (DAPs) were identified (Table 1, Figure 1). Fifteen DAPs were increased in *S. aureus* PJI compared to NIAF samples, including lactotransferrin (LTF) [log_2_fold-change = 5.2], lipocalin-2 (LCN2) [log_2_fold-change = 5.0], myeloperoxidase (MPO) [log_2_fold-change = 4.9], calprotectin-A9 (S100A9 subunit) [log_2_fold-change = 4.5], calprotectin-A8 (S100A8 subunit) [log_2_fold-change = 4.0], cathepsin G (CTSG) [log_2_fold-change = 4.0], neutrophil elastase (ELANE) [log_2_fold-change = 3.8], eosinophil cationic protein (RNASE3) [log_2_fold-change = 3.6], endoplasmic reticulum to nucleus signaling 1 (ERN1) [log_2_fold-change = 3.2], matrix metalloproteinase-9 (MMP9) [log_2_fold-change = 2.9], lysozyme C (LYZ) [log_2_fold-change = 2.7], haptoglobin (HP) [log_2_fold-change = 2.6], lamin-B1 (LMNB1) [log_2_fold-change = 2.5], glycogen phosphorylase, liver form (PYGL) [log_2_fold-change = 2.5], leucine-rich alpha-2-glycoprotein (LRG1) [log_2_fold-change = 2.2] (Table 1).

**Table 1.** Proteins differentially abundant in *Staphylococcus aureus* periprosthetic joint infection (PJI) *versus* non-infectious arthroplasty failure (NIAF) sonicate fluid samples by liquid chromatography-tandem mass spectrometry (LC-MS/MS) analysis. Proteins with increased and decreased abundance in *S. aureus* PJI compared to NIAF samples are unshaded and shaded, respectively. Statistical significance was determined using 2-sample t-test p-values ≤0.05 and log_2_fold-change values ≥2 or ≤-2. Data depicted are for NIAF (n=4) and *S. aureus* PJI (n=4) sonicate fluid samples.

| Protein description | Gene symbol | p-value | $\log_2$ Fold-change |
| --- | --- | --- | --- |
| Lactotransferrin | LTF | 0.0061 | 5.198 |
| Lipocalin-2 | LCN2 | 0.0032 | 5.001 |
| Myeloperoxidase | MPO | 0.0084 | 4.896 |
| Calprotectin (A9 subunit) | S100A9 | 0.0003 | 4.531 |
| Cathepsin G | CTSG | 0.0177 | 4.037 |
| Calprotectin (A8 subunit) | S100A8 | 0.0119 | 3.951 |
| Neutrophil elastase | ELANE | 0.0128 | 3.780 |
| Eosinophil cationic protein | RNASE3 | 0.0154 | 3.629 |
| Endoplasmic reticulum to nucleus signaling 1 | ERN1 | 0.0062 | 3.169 |
| Matrix metalloproteinase-9 | MMP9 | 0.0151 | 2.933 |
| Lysozyme C | LYZ | 0.0049 | 2.699 |
| Haptoglobin | HP | 0.0169 | 2.598 |
| Lamin-B1 | LMNB1 | 0.0007 | 2.492 |
| Glycogen phosphorylase, liver form | PYGL | 0.0059 | 2.471 |
| Leucine-rich alpha-2-glycoprotein | LRG1 | 0.0067 | 2.169 |
| Cathepsin D | CTSD | 0.0385 | -2.007 |
| Fatty acid-binding protein, heart | FABP3 | 0.0197 | -2.031 |
| Fructose-1,6-bisphosphatase 1 | FBP1 | 0.0218 | -2.039 |
| Fatty acid-binding protein, epidermal | FABP5 | 0.0222 | -2.063 |
| Collagen alpha-2(I) chain | COL1A2 | 0.0035 | -2.085 |
| Early endosome antigen 1 | EEA1 | 0.0360 | -2.111 |
| Heat shock protein beta-1 | HSPB1 | 0.0118 | -2.127 |
| Dihydrolipoamide S-succinyltransferase | DLST | 0.0256 | -2.304 |
| Branched-chain-amino-acid aminotransferase | BCAT1 | 0.0151 | -2.315 |
| Annexin A2 | ANXA2 | 0.0015 | -2.363 |
| Serpin B6 | SERPINB6 | 0.0439 | -2.384 |
| CD44 antigen | CD44 | 0.0020 | -2.451 |
| Dermcidin | DCD | 0.0435 | -2.458 |
| Ribonuclease pancreatic | RNASE1 | 0.0184 | -2.541 |
| Proteoglycan 4 | PRG4 | 0.0037 | -2.563 |
| Beta-hexosaminidase subunit beta | HEXB | 0.0494 | -2.662 |
| Osteopontin | SPP1 | 0.0099 | -2.690 |
| IFI30 lysosomal thiol reductase | IFI30 | 0.0237 | -2.798 |
| Cell surface glycoprotein MUC18 | MCAM | 0.0221 | -4.240 |
| Cartilage acidic protein 1 | CRTAC1 | 0.0025 | -5.297 |

**Figure 1.**
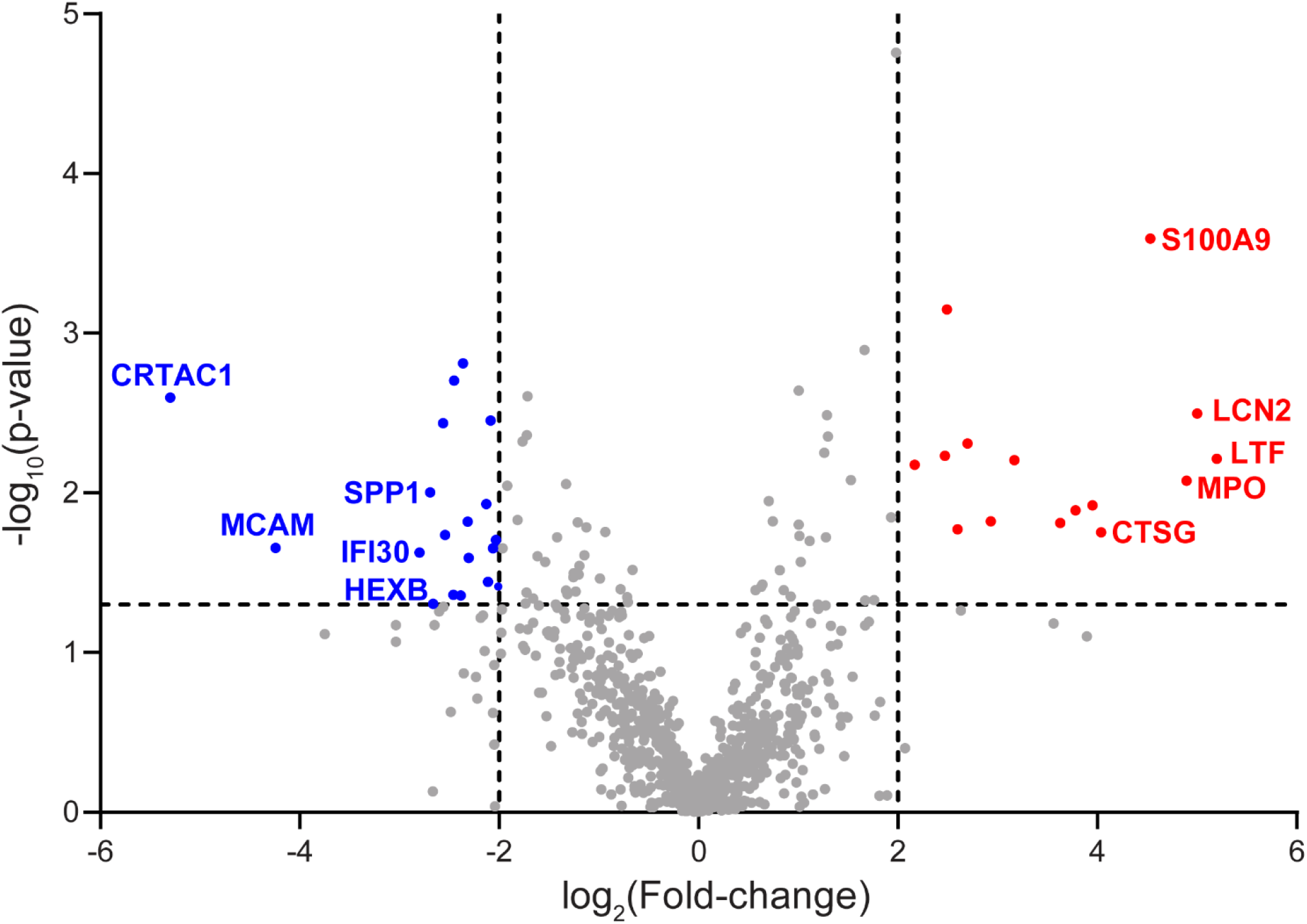
Differential analysis of proteins identified in sonicate fluids from periprosthetic joint infection (PJI) and non-infectious arthroplasty failure (NIAF) using liquid chromatography–tandem mass spectrometry analysis. Volcano plot showing statistically significant proteins differentially abundant in sonicate fluid from PJI cases. Proteins with increased and decreased abundance were indicated in red and blue, respectively. The top five DAPs are labelled. Vertical dashed lines designate log_2_Fold-change value of ±2-fold and the horizontal dashed line designates a p-value = 0.05. Statistical significance was determined using 2-sample t-test with multiple hypothesis correction (p-values ≤0.05 and log_2_fold-change values ≥2 or ≤-2). Data depicted are of NIAF (n=4) and *S. aureus* PJI (n=4) sonicate fluid samples.

Twenty of the 35 DAPs were decreased in *S. aureus* PJI compared to NIAF sonicate fluid samples (Table 1, Figure 1), including cartilage acidic protein 1 (CRTAC1) [log_2_fold-change = - 5.3], cell surface glycoprotein MUC18 (MCAM) [log_2_fold-change = -4.2], IFI30 lysosomal thiol reductase (IFI30) [log_2_fold-change = -2.8], osteopontin (SPP1) [log_2_fold-change = -2.7], beta-hexosaminidase subunit beta (HEXB) [log_2_fold-change = -2.6], proteoglycan 4 (PRG4) [log_2_fold-change = -2.6], ribonuclease pancreatic (RNASE1) [log_2_fold-change = -2.5], dermcidin (DCD) [log_2_fold-change = -2.5], CD44 antigen (CD44) [log_2_fold-change = -2.5], annexin A2 (ANXA2) [log_2_fold-change = -2.4], serpin B6 (SERPINB6) [log_2_fold-change = -2.4], branched-chain-amino-acid aminotransferase (BCAT1) [log_2_fold-change = -2.3], dihydrolipoamide S-succinyltransferase (DLST) [log_2_fold-change = -2.3], heat shock protein beta-1 (HSPB1) [log_2_fold-change = -2.1], early endosome antigen 1 (EEA1) [log_2_fold-change = -2.1], collagen alpha-2(I) chain (COL1A2) [log_2_fold-change = -2.1], fatty acid-binding protein, epidermal (FABP5) [log_2_fold-change = -2.1], fructose-1,6-bisphosphatase 1 (FBP1) [log_2_fold-change = - 2.0], fatty acid-binding protein, heart (FABP3) [log_2_fold-change = -2.0], cathepsin D (CTSD) [log_2_fold-change = -2.0].

Unsupervised clustering analysis of the DAPs showed distinct pattern of expression between *S. aureus* PJI and NIAF sonicate fluid samples. (Figure 2A). Differential clustering was also observed via principal component analysis, where *S. aureus* PJI and NIAF sonicate fluid samples were separated along dimension 1, accounting for 81.8% of the total variation of the dataset (Figure 2B).

**Figure 2.**
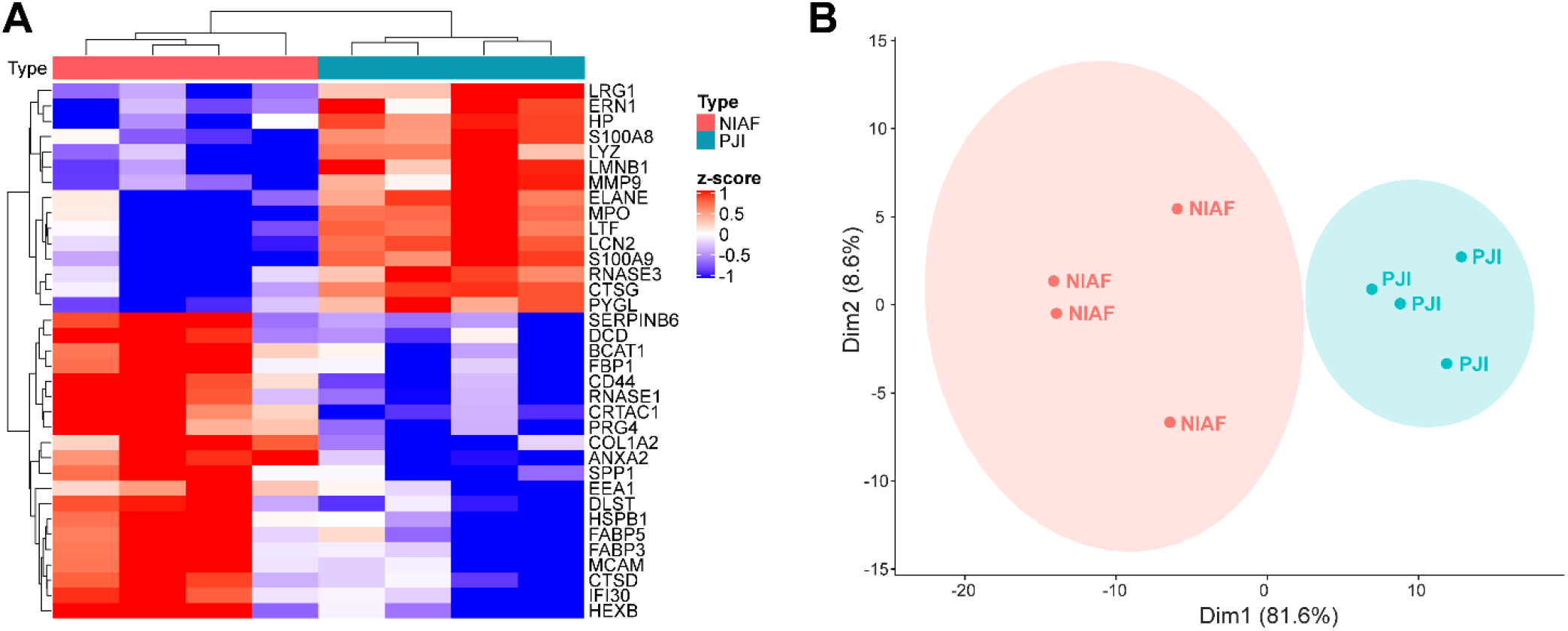
Differential abundant proteins clearly segregate sonicate fluid samples with *Staphylococcus aureus* periprosthetic joint infection (PJI) from non-infectious arthroplasty failure (NIAF). (A) Heatmap visualization of protein expression z-scores in NIAF and *S. aureus* PJI samples, with differentially abundant proteins and patient samples clustered. (B) Principal component analysis of NIAF and *S. aureus* PJI samples, with ellipses corresponding with 85% confidence intervals. Statistical significance was determined using 2-sample t-test with multiple hypothesis correction (p-values ≤0.05 and log_2_fold-change values ≥2 or ≤-2). Data depicted are of NIAF (n=4) and *S. aureus* PJI (n=4) sonicate fluid samples.

### 3.2 Gene ontology analyses of differentially abundant proteins

To better understand the functional outcome of sonicate fluid DAPs in PJI *versus* NIAF, gene ontology analysis was conducted ^[53-55]^. Unsurprisingly, biological processes related to neutrophil antimicrobial activity, such as activation and degranulation, were increased in *S. aureus* PJI compared to NIAF sonicate fluid samples. Overall antimicrobial responses against bacteria and fungi were also increased in *S. aureus* PJI (Figure 3A). Next, molecular functions enriched among DAPs were examined. Inflammatory pathways, such as those related to peptidase activity and RAGE receptor binding were increased in *S. aureus* PJI samples. Metal ion-binding pathways were also increased in *S. aureus* PJI samples. In fact, many of the DAPs most increased in *S. aureus* PJI, such as LTF, LCN2, and S100A8/A9, are known metal ion sequestration proteins reported to be important for the antimicrobial nutritional immunity response in other infection types ^[56-58]^. Though increases of metal ion binding proteins have recently been shown to be clinical biomarkers of PJI biomarkers ^[27,28,30,31,59,60]^, the functional role of nutritional immunity during PJI is an area needing further investigation.

**Figure 3.**
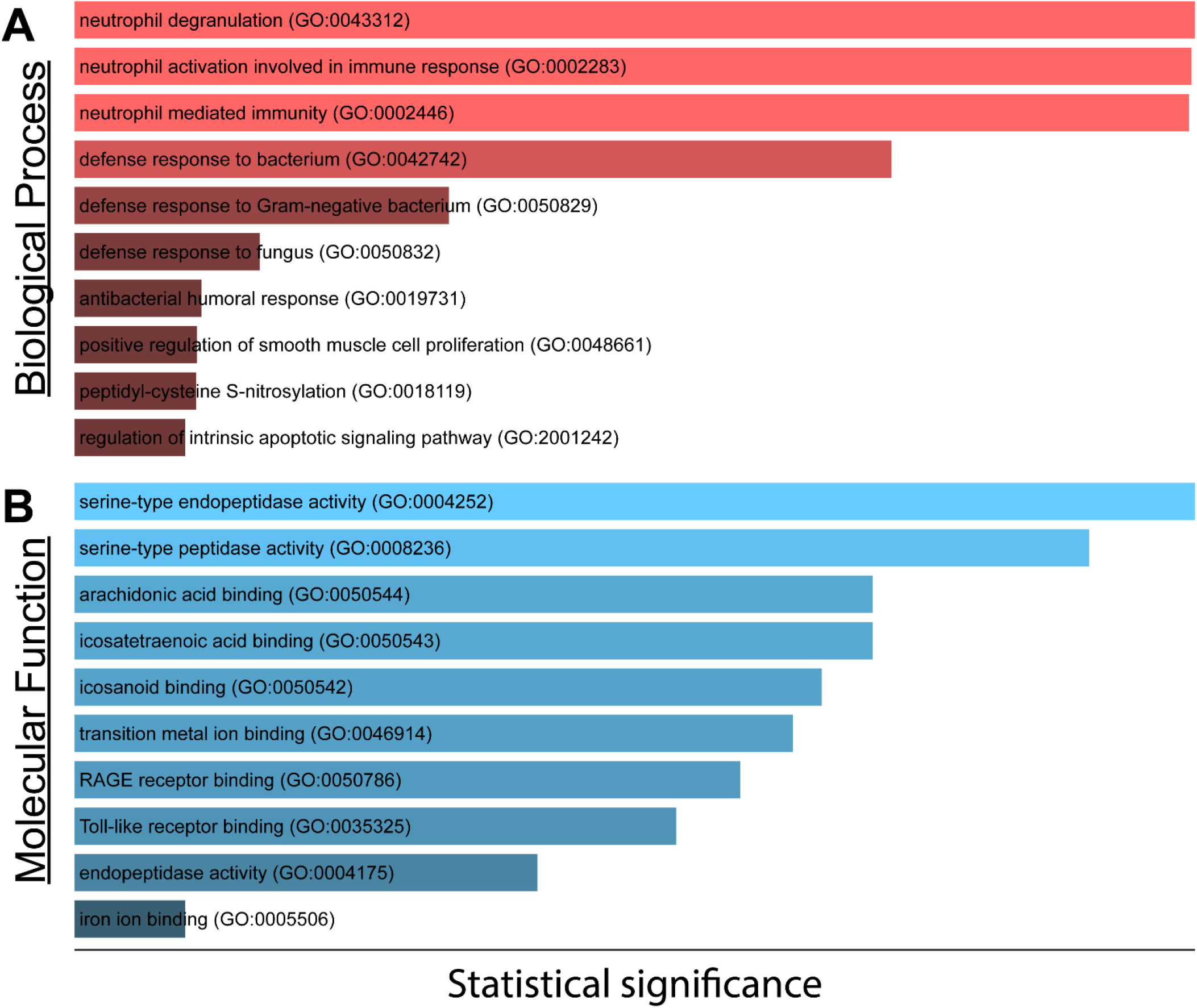
Gene ontology analysis of proteins differentially abundant in *S. aureus* periprosthetic joint infection (PJI) *versus* non-infectious arthroplasty failure (NIAF) sonicate fluid samples. (A) Biological processes and (B) Molecular functions enriched using the Gene Ontology 2021 knowledgebase. Gene ontology terms listed with those most statistically significant on top. Data depicted are of NIAF (n=4) and *S. aureus* PJI (n=4) sonicate fluid samples.

### 3.3 Further validation of differentially abundant proteins

While the mass-spectrometry-based proteomic profiling described here identified DAPs and differentiated *S. aureus* and NIAF sonicate fluid samples, the sample size was low, and further validation was necessary. To do so, results were compared to previous transcriptomic and proteomic analyses of sonicate fluid samples conducted by our group. Transcriptomic analysis of eight *S. aureus* PJI and forty NIAF sonicate fluid samples, including the eight that underwent LC-MS/MS here, has been recently reported ^[40]^. Of the thirty-five DAPs found via LC-MS/MS, 18 had differentially expressed transcripts via RNA-sequencing. In all but one recapitulated case, whether the target was up- or downregulated in *S. aureus* PJI compared to NIAF remained consistent. MMP9 was found to have increased protein abundance but decreased transcriptomic abundance in *S. aureus* PJI compared to NIAF samples.

Proteomic profiling of 200 sonicate fluid samples, including the eight that underwent LC-MS/MS, was recently reported using the proximity extension assay (PEA) platform from Olink Proteomics (Uppsala, Sweden) ^[34,61]^. PEA showed that, of the 92 proteins included in the Olink Inflammation Panel, 37 DAPs were found when comparing PJI and NIAF samples ^[34]^. Interestingly, there was no overlap across the 37 DAPs identified through PEA platform to the 35 DAPs identified by LC-MS/MS analysis. This lack of overlap is explained by specific characteristics of each platform. PEA is a targeted method that uses antibodies conjugated to oligonucleotides and PCR amplification to process and quantify proteins, allowing for assessment of low-concentration proteins. However, this analysis is limited to detection to the specific protein targets included in the panel ^[61]^. In contrast, LC-MS/MS is an unbiased approach, which can quantify proteins in a global fashion that are above the detection limit ^[62]^. Thus, a targeted approach like PEA is complementary to an LC-MS/MS approach. Thus, combining LC-MS/MS with PEA provides a holistic proteomic profile for protein biomarker discovery.

Lastly, results were compared to those found by Li et al. which analyzed 51 PJI and 66 non-PJI synovial fluid samples identified by LC-MS/MS ^[41]^. Of the 35 DAPs found in sonicate fluid, 15, including LTF, MPO, S100A9, CTSG, S100A8, ELANE, MMP9, HP, PYGL, CTSD, FABP5, BCAT1, PRG4, HEXB, and CRTAC1, were also found in synovial fluid. Proteomic analysis of synovial fluid revealed 281 DAPs, a higher number than identified herein in sonicate fluid, with the caveat that more patients and bacterial species causing PJI were analyzed in the synovial fluid study. That proteomic analyses of sonicate and synovial fluid were not fully concordant affirms the potential value of interrogating both sample types, though ideally synovial and sonicate fluids from the same patients, collected at the same time and analyzed using the same methods, should be analyzed.

## 4 CONCLUDING REMARKS

Overall, results from this study indicate that proteomic profiling of sonicate fluid using mass spectrometry-based proteomic analysis differentiates *S. aureus* PJI and NIAF samples. This study supports the concept of conducting larger and more thorough future proteomic profiling studies using combinational analyses and complementary sample-types.

## Data Availability

The mass spectrometry proteomics data have been deposited to the Proteome Xchange Consortium via the PRIDE partner repository with the dataset identifier PXD038928.

## ABBREVIATIONS

PJI: periprosthetic joint infection
NIAF: non-infectious arthroplasty failure
DAPs: differentially abundant proteins
LC-MS/MS: liquid chromatography-tandem mass spectrometry
CCL20: chemokine ligand 20
SARS-CoV-2: severe acute respiratory syndrome coronavirus 2
LTF: lactotransferrin
LCN2: lipocalin-2
MPO: myeloperoxidase
S100A9: calprotectin-A9
S100A8: calprotectin-A8
CTSG: cathepsin G
ELANE: neutrophil elastase
RNASE3: eosinophil cationic protein
ERN1: endoplasmic reticulum to nucleus signaling 1
MMP9: matrix metalloproteinase-9
LYZ: lysozyme C
HP: haptoglobin
LMNB1: lamin-B1
PYGL: glycogen phosphorylase, liver form
LRG1: leucine-rich alpha-2-glycoprotein
CRTAC1: cartilage acidic protein 1
MCAM: cell surface glycoprotein MUC18
IFI30: IFI30 lysosomal thiol reductase
SPP1: osteopontin
HEXB: beta-hexosaminidase subunit beta
PRG4: proteoglycan 4
RNASE1: ribonuclease pancreatic
DCD: dermcidin
CD44: CD44 antigen
ANXA2: annexin A2
SERPINB6: serpin B6
BCAT1: branched-chain-amino-acid aminotransferase
DLST: dihydrolipoamide S-succinyltransferase
HSPB1: heat shock protein beta-1
EEA1: early endosome antigen 1
COL1A2: collagen alpha-2(I) chain
FABP5: fatty acid-binding protein, epidermal
FBP1: fructose-1,6-bisphosphatase 1
FABP3: fatty acid-binding protein, heart
CTSD: cathepsin D
PEA: proximity extension assay
TCEP: tris(2-carboxyethyl)phosphine
FDR: false discovery rate

## STATEMENT OF CLINICAL SIGNIFICANCE

Periprosthetic joint infection (PJI) is a major complication of joint arthroplasty. There is no perfect assay for differentiating PJI from non-infectious arthroplasty failure (NIAF). Although some studies have recently employed ‘omics technologies, further work is needed to discover novel and sensitive biomarkers and increase accuracy. A comprehensive proteomic analysis of sonicate fluid from PJI, a specimen derived from sonication of resected implants to sample their surfaces, is not yet reported.

## ACKNOWLEDGEMENTS

This research was supported by the National Institute of Arthritis and Musculoskeletal and Skin Diseases of the National Institutes of Health under Award Number NIH R01 AR056647. CF was supported by the Mayo Clinic Graduate School of Biomedical Sciences and the Ph.D. Training Grant in Basic Immunology (NIAID T32 AI07425-25). The content is solely the responsibility of the authors and does not necessarily represent the official views of the NIH. We would like to acknowledge Suzannah Schmidt-Malan, M.S., Melissa Karau, M.S., and the rest of the Mayo Clinic Infectious Diseases Research Laboratory for accessioning and maintaining the sonicate fluid biobank.

## PATIENT CONSENT STATEMENT

The study was approved by the Mayo Clinic Institutional Review Board (#09-000808).

## AUTHORS’ CONTRIBUTIONS

CF’s roles were conceptualization, data curation, formal analysis, investigation, visualization, writing – original draft, and writing – review & editing. KM’s roles were mass spectrometry analysis, data analysis and visualization, and writing – review & editing. KG’s roles were project administration, supervision, and writing – review & editing. MA’s roles were resource provision and writing – review & editing. AP’s roles were resource provision, supervision, and writing – review & editing. RP’s roles were conceptualization, funding acquisition, project administration, resource provision, supervision, and writing – review & editing. All authors read and approved the final manuscript.

